# The HIV-Associated Neurocognitive Disorders in Zambia (HANDZ) Study: Protocol of a research program in pediatric HIV in sub-Saharan Africa

**DOI:** 10.1101/19003590

**Authors:** Heather R. Adams, Sylvia Mwanza-Kabaghe, Esau G. Mbewe, Pelekelo P. Kabundula, Michael J. Potchen, Sanjay Maggirwar, Brent A. Johnson, Giovanni Schifitto, Harris A. Gelbard, Gretchen L. Birbeck, David R. Bearden

**Author notes:** Financial Disclosure: All authors have no financial relationships relevant to this article to disclose. Conflict of Interest: All authors have no relevant conflicts of interest to disclose. Clinical Trial Registration: Not applicable. Contributors Statement Page: Heather Adams: Dr. Adams conceptualized and designed the study, drafted and revised the manuscript, and approved the final manuscript as submitted., David Bearden: Dr. Bearden conceptualized and designed the study, assisted in drafting and revising the manuscript, and approved the final manuscript as submitted., All other authors assisted in conceptualizing and designing the study, collected data or advised on the process of data collection, assisted in drafting and revising the manuscript, and approved the final manuscript as submitted.

## Abstract

Approximately 10% of youth in sub-Saharan Africa are infected with the Human Immunodeficiency Virus. In Zambia, it is estimated that over 72,000 children have HIV infection, and despite access to combination antiretroviral therapy, many will experience HIV-associated neurocognitive deficits (HAND) encompassing cognitive and psychiatric sequelae such as global intellectual delay, executive dysfunction, and depressed mood. However, little is known about the neurocognitive profile of such children, the long-term outcomes and impacts of HAND, or the predictors and risk factors for HAND-related impairment. We have initiated the first-ever prospective, longitudinal study of neurocognition in children with HIV-infection in Zambia. Our overarching study goals are to validate cognitive and psychiatric testing tools in children with HIV infection in Zambia, and to determine if inflammatory biomarkers and brain imaging can prospectively identify children at high risk of developing HAND. This article outlines the study methods, highlights several challenges encountered in the initiation of the study, and offers solutions to these challenges.

## INTRODUCTION

Approximately 3.4 million children and adolescents in sub-Saharan Africa are infected with the Human Immunodeficiency Virus (HIV), amounting to about 10% of all persons in the region living with HIV or AIDS. In Zambia, an estimated 72,000 children (age 0-14 years) are HIV infected, representing about 1% of the population within that age group. Approximately 64% of children living with HIV in Zambia receive combination antiretroviral therapy (cART) and the majority now begin treatment early in life^1,2^. Despite access to cART, children infected with HIV in Zambia and elsewhere in the region remain at risk for developmental delay and cognitive impairment. Little is yet known about the cognitive profile of HIV-infected children in Zambia, but studies of HIV-positive children living in other sub-Saharan nations reveal delayed attainment of developmental milestones and poor performance on tests of general cognitive abilities (e.g., IQ) as well as neuropsychological tasks of attention, processing speed, executive function, learning and memory, even with early initiation of cART^3-5^. Among children with HIV who obtain scores in the normal (average) range on tests of general intelligence, HIV-positive children perform less well on tests of these specific neurocognitive skills, in comparison to demographically matched control subjects who are HIV-unexposed^6^.

While cART has demonstrated effectiveness in suppressing viral load, preserving immune function and improving general medical health in children with HIV, an estimated 20%-50% of children treated with cART may nonetheless develop cognitive deficits^7^. There are several possible explanations for this including poor penetration of some antiretroviral drugs (ARVs) into the CNS, sustained CNS inflammation despite viral suppression, neurotoxicity of some ARVs and the impact of comorbidities on cognitive functioning. Additionally, children and adolescents with HIV may be at increased risk for psychiatric disorders such as anxiety and depression both due to psychosocial consequences of having HIV, and the pathogenic effects of HIV on brain development that may predispose to psychiatric disease. Mood dysfunction may also impact cognitive test performance. Finally, in low-resource settings such as Zambia, co-varying adverse environmental exposures including malnutrition, disrupted schooling, psychosocial stressors, and lower birthweight among perinatally-exposed children born to HIV-infected mothers may also impact cognitive outcomes^8^.

Because little is known about the complex influences on neurocognitive and psychiatric function in HIV-infected Zambian children or the natural history of cognitive development in his population over time, we have established a three-year prospective, longitudinal study of cognitive development children and young adults ages 3-19 years old with and without HIV-infection living in Zambia. This study includes a longitudinal cohort component evaluating differences between HIV-infected and HIV-exposed uninfected (HEU) controls, and a case control component evaluating risk factors for cognitive impairment and cognitive decline. The objective of this article is to describe the study aims and methods, and discuss several challenges and solutions related to the design and implementation of this study.

## STUDY AIMS AND OBJECTIVES

The overarching goals of the study are to validate cognitive and psychiatric testing tools in children with HIV in Zambia, and to determine whether inflammatory biomarkers and brain imaging can prospectively identify children at high risk of developing cognitive impairment. The specific research aims are: 1) To identify culturally appropriate tools for the assessment of cognitive impairment and psychiatric comorbidities in children and adolescents with HIV in Zambia; 2) To determine the prevalence and annual incidence of cognitive impairment, major depression, and generalized anxiety among children and adolescents with HIV in Zambia; 3) To identify serum biomarkers associated with cognitive impairment in children and adolescents with HIV in Zambia; 4) To identify imaging findings associated with cognitive impairment in children and adolescents with HIV in Zambia, and 5) To develop a predictive model of incident cognitive decline in children and adolescents with HIV. Ultimately, our long-term objectives are to build capacity for future studies of HIV associated neurocognitive disease in Zambian children, and in doing so, to set the stage for future clinical trials to treat or prevent the neuropsychiatric complications of HIV infection in Zambian youth.

## STUDY ORGANIZATION

### Overview

The University of Rochester Medical Center (URMC) in Rochester, NY USA is the academic site and coordinating center for the study (PI: Bearden); all study activities are conducted at the Paediatric HIV/AIDS Center of Excellence in Lusaka, the capital city of Zambia. The PI (DRB) works in Zambia at the study site for approximately 3 months per year, and meets weekly via video or phone-conference with the local study team when not in the country. A URMC-based Neuropsychologist (HRA) developed the neurocognitive assessment battery in collaboration with the Zambian investigators and travels to Zambia approximately 1-2 times per year for study training, data audits, and informal didactic instruction on topics relevant to neurocognitive assessment in children. A neuroradiologist (MP) with joint appointments at URMC and the Lusaka Apex Medical University in Zambia assisted in designing the imaging protocol, developed the database for the imaging portion of the study, and reviewed all Magnetic Resonance Imaging (MRI) studies. Zambian study staff include a supervising PhD level Psychologist (SMK), two Master’s level graduate students in Educational Psychology who conduct neurocognitive assessments (EG, PK), and a study nurse, study clinical officer, research assistant, and lab technologist.

### Study Site and Participants

Study visits occur at the University Teaching Hospital (UTH) Pediatric HIV/AIDS Centre of Excellence (PCOE), the major HIV referral center in Zambia. Recruitment began in November 2017. At completion of the first year of the study, our pre-planned target sample of 200 children and adolescents with perinatally-acquired HIV infection (cases) and 200 HIV-exposed uninfected youth (controls, HEU) aged 8-17 was enrolled. In April 2019, inclusion criteria were expanded to children age 3-7 years old with a corresponding sample size increase within that age range to add 200 additional subjects (n=100 HIV-infected, N=100 HEU). The rationale for expanding recruitment to this additional cohort of younger subjects is discussed below.

HIV-infected youth are recruited at routine outpatient medication refill visits to PCOE. Stratified sampling is used to ensure participants in each group are approximately equally distributed across the age range, maintain an equal sex distribution within and between groups, and to recruit HEU subjects (via a community health social worker) from the same neighborhoods in which HIV-infected subjects reside. Imbalances in potential confounding variables are adjusted for in the statistical analysis (see details in analysis plan, below).

### Inclusion / Exclusion Criteria

Eligible subjects are males or females between 3-19 years old at time of enrollment with at least one living parent available to give consent and participate in the study. Subjects with HIV-positive status confirmed by Western blot or DNA PCR and current treatment with cART are eligible to participate as cases. We required cART initiation at least one year prior to enrollment to ensure stable disease and treatment. Subjects with maternal exposure during pregnancy and HIV-negative status confirmed by immunoassay are eligible to participate as controls. For both groups, exclusion criteria include: parent unable or unwilling to provide permission for the child’s participation; history of central nervous system infection; chronic or acute medical or psychiatric conditions other than HIV that could potentially impact study participation or results including psychosis, malignancy, or chronic liver or renal failure; currently taking immunosuppressive drugs; sensory, language, or motor impairment that in the opinion of the study physician would impede ability to participate in the study or complete study measures (described below); HIV infection known to be secondary to sexual contact or blood transfusion; inability of child to understand and communicate basic concepts in English. We also excluded individuals with plans to leave Lusaka City or transfer care away from PCOE during the study.

#### Neuroimaging sub-study

From the overall sample, a target sample size of n=34 children in each group is also recruited to participate in a neuroimaging sub-study (Study Aim 4) to obtain MRI and magnetic resonance angiography (MRA) of the brain. Recruitment for this sub-study has been completed for the HIV infected group and continues for the HEU group. Additional exclusion criteria for this imaging sub-study are: younger than 8 years old, any medical or psychiatric condition that would preclude a non-sedated MRI/MRA including severe claustrophobia or in-dwelling metal objects; pregnancy. A urine pregnancy test is administered to female subjects immediately prior to MRI scan; those with a positive test are excluded from this sub-study.

### Subject Remuneration and Informed Consent

Remuneration is provided in accordance with local standards to offset lost time/travel to the study site. Parental permission is obtained from the parents of all children enrolling in the study and assent is obtained from children age 12 years and older. Per local standards, subjects age 16-17 years’ old who attend visits by themselves complete an adult informed consent process and any subject providing assent (with parental permission) who becomes 18 years old during the study is re-consented as an adult. English is the official language of Zambia and documents such as consent forms are expected to be provided in this language. Among parents who cannot read, the study coordinator performs the consent process in the parent’s preferred language. For parents who cannot write their own name, consent is documented via thumbprint. Adolescent/young adult subjects who consent to their own participation (age 16 years and older) must be capable of doing so in English, as their ability to converse in this language is a requirement for study participation (due to the English language-based neurocognitive assessment). All study activities are dually reviewed and approved by the Institutional Review Boards (IRBs) of the University of Rochester (Research Subjects Review Board protocol #00068985) and the University of Zambia which is the reviewing IRB for investigations conducted at PCOE-UTH (reference #004-08-17), as well as by the Zambian National Health Research Authority.

## STUDY VISITS, METHODS, PROCEDURES

Subjects complete comprehensive annual study visits and brief interim study visits on a quarterly basis. At annual visits, subjects undergo comprehensive assessment of mood, behavior, and neurocognitive abilities, medication adherence checks, phlebotomy, and review of interim medical and social history. The baseline annual visit also includes a detailed review of sociodemographic background and child developmental and medical history. Quarterly visits include a brief assessment of cognition and mood, medication adherence checks, and review of interim medical and social history. **Table 1** shows the schedule of activities, organized by study visit.

**Table 1.**
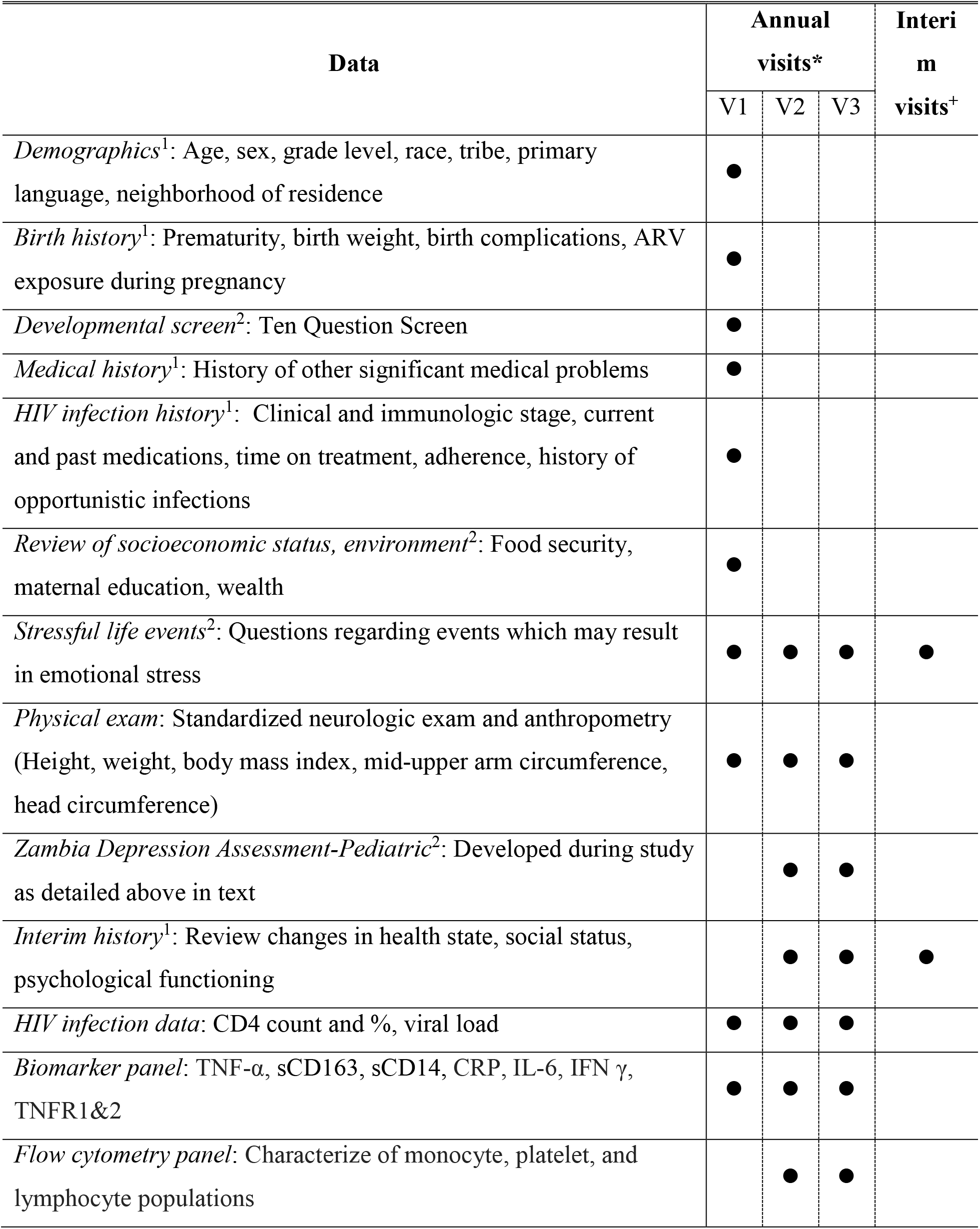
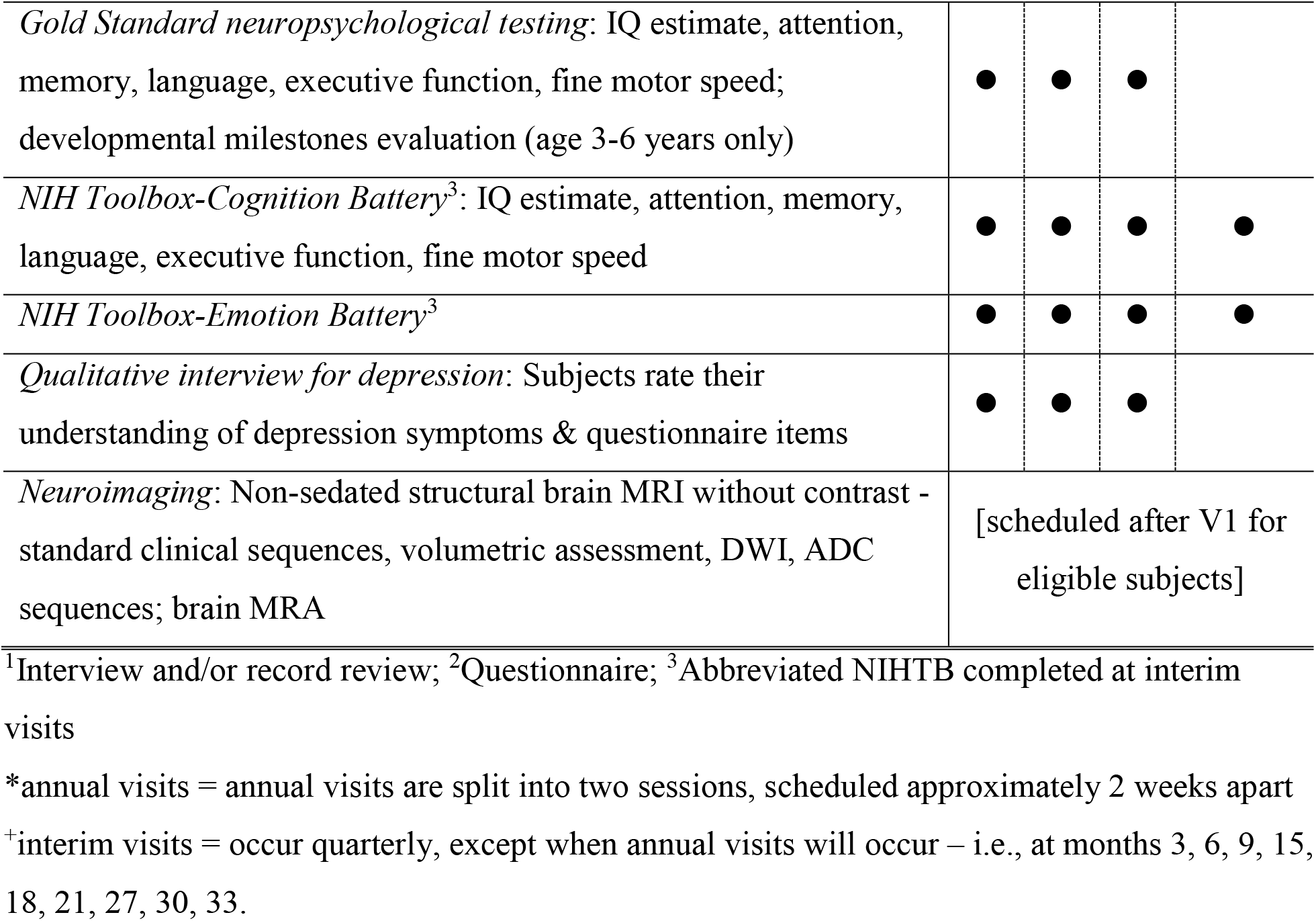
HANDZ Study Visits - Schedule of Activities.

### Subjects’ background information

#### Socioeconomic Status (SES) and Demographic Information

SES is evaluated using a measure adapted from the Multiple Indicator Cluster Survey-5 (MICS5, http://mics.unicef.org/tools). This tool was adapted for the current study by eliminating questions not relevant in a predominantly urban setting. The questionnaire evaluates access to running water and electricity, toilet facilities, food security, ownership of household items that are markers of higher income and access to resources (e.g., radio, mobile phone, television). The local staff conducting this interview also rates the subject’s SES, relative to the typical patient seen at PCOE. Social and demographic variables include details about the subjects’ neighborhood and household including primary (and other) languages spoken in the home, who his/her primary caretaker is, school information, parental educational level and occupation, and monthly household income.

#### Developmental Screening

The 10-Question Screen^9^ evaluates developmental history and is an established screening tool for use in low-resource settings. Questions address delays in meeting motor and language milestones, any hearing or vision problems, cognitive delay, motor impairment, and seizures. Through parental interview and/or chart review, information is also obtained about the subject’s prenatal and birth history, including whether prenatal care was received, maternal alcohol or tobacco use during pregnancy, any complications during the mother’s pregnancy or delivery, and during the perinatal and postnatal periods. For children with HIV-infection, information is also collected about maternal HIV health history, including ARV treatment and maternal CD4 count during pregnancy and delivery, and maternal WHO Clinical stage utilizing both parental interview and maternal medical chart. The developmental screening is completed at the baseline visit only.

#### HIV History (Parent and Child)

For HIV-positive subjects, the child’s HIV-infection related history is obtained via parent interview and through chart review. At the initial (baseline) evaluation the following HIV history variables are collected:

- Treatment history: age of cART initiation, current and past ARV medications, any treatment interruptions lasting longer than 2 weeks
- Infection severity: Most recent and lowest recorded CD4 count and percentage, worst recorded WHO Clinical Stage and reason (e.g., opportunistic infection [OI], any past OIs, other HIV complications including those necessitating hospitalization.

At all annual and interim (quarterly) follow-up visits, this history is again reviewed and updated, and any lapses in cART of 3 days or more are recorded and investigated.

#### Physical Exam

At all annual visits subjects complete a physical exam including anthropometry (height, weight, body mass index, mid-upper arm circumference, head circumference) and a standard neurologic exam.

### Study Aim 1: Establish Culturally Appropriate Tools to Assess Cognition and Mood in HIV-Infected Youth in Zambia

The first study aim is to identify culturally appropriate tools to assess cognition (Aim 1a) and mood (Aim 1b) in Zambian youth. An additional goal is to evaluate assessment strategies that are feasible in this low-resource setting, particularly with respect to cognitive assessment. Even in Zambia’s capital city of Lusaka, there are few individuals qualified to administer or interpret neuropsychological evaluations, and access to tests is limited by cost and distance. The NIH Toolbox (NIHTB) for Assessment of Neurologic and Behavioral Function^10^ offers a potential solution to these challenges. It is a tablet computer-based battery of open-access tests of cognitive, sensory, neurologic and behavioral function. It has been validated in children and adolescents in the United States^11-13^, and the PI (Bearden) has previously successfully utilized the Toolbox in pilot studies with children in Botswana and Liberia (*unpublished data*). NIHTB measures are portable and designed to be administered by a trained study coordinator or nurse.

#### Aim 1a. Culturally Appropriate Tools to Assess Cognition

For Study Aim 1a, we will determine if the NIHTB-Cognition Battery (NIHTB-CB) can be used to evaluate cognitive impairment and change in neurocognitive function over time in children and adolescents with HIV-infection, in comparison to HEU control subjects. Performance on the NIHTB-CB tasks will be compared to a gold-standard neuropsychological assessment, using traditional paper-and pencil measures. Each assessment (NIHTB-CB and gold-standard) includes tasks to assess attention, memory, executive function, processing speed and fine motor function, domains known to be at risk for impairment in children with HIV infection. The NIHTB-CB is administered by a study nurse or coordinator; the traditional neuropsychological battery is administered by Master’s level Psychologists (PK; EM) pursuing their PhD in Educational Psychology at the University of Zambia. They were trained by a pediatric neuropsychologist (HRA) and are supervised locally by a PhD-level educational psychologist (SMK) with experience in research evaluations in Zambia. After completing the NIHTB-CB subjects are asked to note any difficulty understanding how to complete each task, and to describe, in their own words, the purpose of each test. The examiner also rates their impression of the child’s comprehension of task instructions and whether the test appeared to measure the intended construct (e.g., sustained attention, or processing speed). **Table 2** presents the neuropsychological assessment tasks and corresponding NIHTB-CB tasks organized by cognitive domain.

**Table 2.**
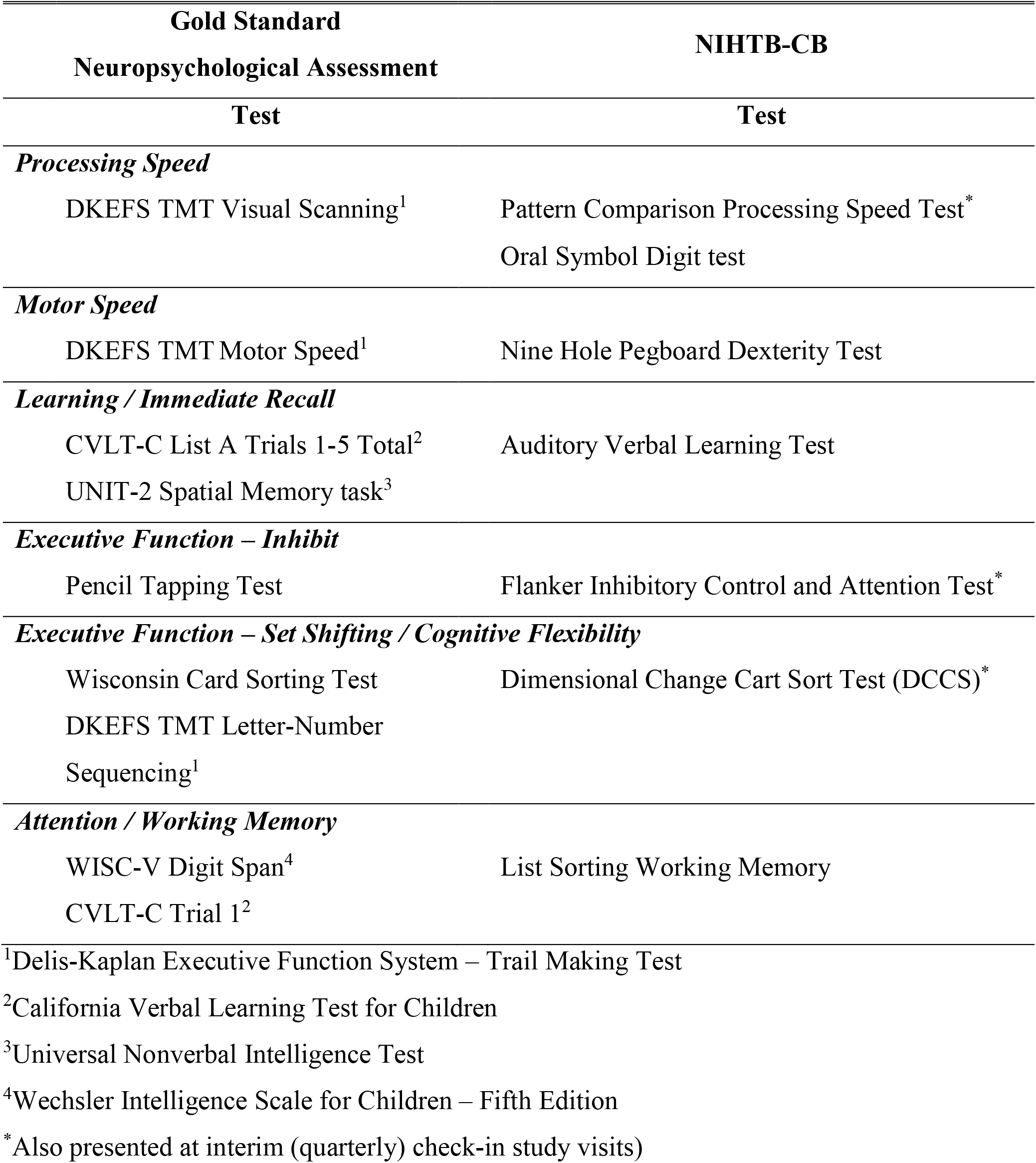
Validation of the NIH Toolbox Cognitive Battery.

The NIHTB-CB and gold-standard assessments are administered at baseline and annual follow-up visits (Years 2, 3), but at separate testing sessions (scheduled approximately 2 weeks apart), to minimize fatigue and interference among similar tasks across the two testing batteries. We plan to identify a NIHTB-CB score that discriminates between children with and without neurocognitive impairment, and evaluate change in this score in relation to the gold standard assessment over a one-year period. The NIHTB-CB is also designed to be repeatable over relatively short re-testing intervals, allowing more frequent monitoring for potential changes in cognitive function. Therefore, at interim (quarterly) study visits, subjects complete an abbreviated NIHTB-CB battery to assess processing speed and executive function, also shown in Table 2.

#### Aim 1b. Culturally Appropriate Tools to Assess Depression

There are presently no standardized, culture-specific tools available to evaluate depressive symptoms among youth in Zambia. It is not known if the clinical triad of cognitive, somatic, and affective symptoms of depression applies to youth in Zambia, or whether other components of the clinical diagnosis related to symptom duration, impairment, or associated features are equally relevant. We also do not know if existing tools to assess mood symptoms in young persons, developed and normed principally with European and North American populations, have adequate face validity and construct validity for use in Zambia. To address these concerns, our study includes a multi-year project to develop a Zambia-specific assessment tool for depression in youth. In Year 1, a panel of local experts (representing fields of psychiatry, psychology, medicine, education, and social anthropology) and study investigators conducted a qualitative review of the relevance and validity of the Diagnostic and Statistical Manual for Mental Disorders – Fifth edition (DSM-5) diagnostic criteria for Major Depressive Disorder (MDD), for evaluating depression in Zambian youth. DSM-5 MDD criteria were rated for face validity, construct relevance, and importance, and we solicited input on additional items for consideration. From this, a pilot version of a new measure, the Zambian Depression Assessment-Pediatric (ZDAP), is being tested in Year 2 of the study. In this pilot phase, subjects complete a cognitive interviewing process regarding comprehension, clarity of wording, and perceived relevance of each item on the ZDAP. We also ask if item content is objectionable and whether the overall length of the measure is acceptable. Revisions to the ZDAP will be made based on this input. Our final phase will be implemented in Year 3 of the overall HANDZ study, during which the revised version of the ZDAP will be administered to each subject at their Year 3 annual visit and a total score based on subject responses will be generated. ZDAP scores will be compared to age- and gender-adjusted T-scores of the Fear and Sadness modules from the NIH Toolbox Emotion Battery to evaluate concurrent validity; and each instrument will be compared to a modified gold standard of expert determination of depressed/not depressed status.

### Study Aim 2a: Determining Prevalence and Incidence of Cognitive Impairment, Depression, and Anxiety in HIV-Infected Youth in Zambia

#### Study Aim 2b: Identifying longitudinal trajectories and risk factors for each outcome described above

As noted above, subjects complete an extensive gold-standard neuropsychological assessment at each annual visit; in addition to providing validation of the NIHTB-CB, this evaluation will support completion of study Aim 2, which is, in part, to determine the prevalence and annual incidence of cognitive impairment among HIV-infected youth in Zambia compared to a HEU control group. As part of the neuropsychological assessment, subjects age ≥6 years old also complete an evaluation of their general cognitive abilities (IQ), using the Universal Nonverbal Intelligence Test-Second edition (UNIT-2)^14^. Subjects age 3-6 years old complete the Malawi Developmental Assessment Tool (MDAT)^15^ to assess attainment of developmental milestones. Though developed and validated for use in Malawi, our selection of the MDAT was guided by concurrent use in another study of neurodevelopmental outcomes of HIV-infected young children in Zambia, to permit comparability across these respective studies. This is discussed further below where we address a challenge (and solution) related to expansion of our sample size age range to include preschool-age children.

In parallel with development of a culturally relevant depression measure for Zambian youth with HIV infection, subjects will also complete existing standardized tools to assess mood, behavior, and social function to ascertain the prevalence of depressed and anxious mood in this population. These assessments are conducted at baseline (Year 1) and annual visits (Year 2, 3) through interviews with study staff and questionnaires. The parent-proxy and child self-report Patient Health Questionnaire, nine item (PHQ-9) Modified form for Adolescents^16,17^ is completed to assess the presence and severity of symptoms of depression. Subjects age 12 years and older also complete the Screen for Child Anxiety and Related Disorders (SCARED)^18-20^, a brief questionnaire to screen for symptoms of Panic disorder, Generalized, Separation, and Social Anxiety disorders, and school avoidance problems. Parents and children also complete the Brief Impairment Scale^21^ (BIS; proxy- and self-report versions, respectively) to assess interpersonal, school, and social function. We have added several items to the School/Work subscale to obtain further detail about any health-related school absences, particularly relevant for our HIV-infected sample of interest. The parent-proxy and child self-report domains from the NIHTB-EB will be compared to the SCARED and BIS. At both comprehensive annual and brief quarterly study visits, subjects also complete a review of recent exposure to events and experiences that might impact mood and daily function, including worsening of physical health, hospitalization of oneself or family members, change in household membership or living arrangements, death of a family member, and experiences of violence or abuse. Finally, at the interim (quarterly) study visits, children complete two modules from the NIHTB-Emotion Battery: Perceived Stress and Sadness. As with the gold-standard neuropsychological assessment, the data obtained from the PHQ-9 and SCARED will help inform prevalence and annual incidence of depression and anxiety symptoms in HIV-infected Zambian youth, compared to HEU controls (Study Aim 2). All measures used in the study to assess cognition, mood, and behavior, are listed in **Table 3**. This table also shows the age ranges for administration of each measure, since some assessments are age-specific.

**Table 3.** HANDZ Neuropsychological Assessment Battery, Mood/Behavior Assessment, and NIH Toolbox Cognitive and Emotion Batteries.

| Measure | Age (years) |  |  |
| --- | --- | --- | --- |
|  | 3-5 | 6-7 | 8-19 |
| *Gold Standard Neuropsychological Assessment* |  |  |  |
| Letter-Number Screening |  | ● | ● |
| Malawi Developmental Assessment Test<sup>1</sup> | ● | ● |  |
| Universal Nonverbal Intelligence Test – Second edition |  | ● | ● |
| Delis Kaplan Executive Function System Trail Making Test<sup>2</sup> |  | ● | ● |
| California Verbal Learning Test for Children |  |  | ● |
| Pencil Tapping Test |  | ● | ● |
| NEPSY-2 Affect Recognition |  | ● | ● |
| NEPSY-2 Verbal Fluency<sup>3</sup> |  | ● | ● |
| Wisconsin Card Sorting Test |  |  | ● |
| WISC-V Digit Span |  | ● | ● |
| *NIH Toolbox Cognition Battery<sup>4</sup>* |  |  |  |
| Pattern Comparison Processing Speed Test |  | ● | ● |
| Flanker Inhibitory Control & Attention Test |  | ● | ● |
| Dimensional Change Card Sort Test |  | ● | ● |
| 9-Hole Pegboard Test |  | ● | ● |
| 4-Meter Walk Gait Speed Test |  | ● | ● |
| List Sorting Working Memory Test |  |  | ● |
| Auditory Verbal Learning Test |  |  | ● |
| Oral Symbol Digit Test |  |  | ● |
| *Mood and Behavior Assessment* |  |  |  |
| Patient Health Questionnaire-9, Youth Self Report |  |  | ● |
| Patient Health Questionnaire-9, Parent Proxy Report |  | ● | ● |
| Behavior Rating Inventory of Executive Function – 2<sup>nd</sup> edition |  | ● | ● |
| Brief Impairment Scale |  |  | ● |
| *NIH Toolbox Emotion Battery<sup>4</sup> - Parent Proxy Measures* |  |  |  |
| Fear |  | ● | ● |
| Perceived Stress |  | ● | ● |
| Sadness |  | ● | ● |
| Positive Parental Relationship |  | ● | ● |
| Self-Efficacy |  | ● | ● |
| Domain Specific Life Satisfaction |  | ● | ● |
| *NIH Toolbox Emotion Battery*<sup>4</sup> - Self Report Measures |  |  |  |
| Emotional Support |  |  | ● |
| Fear |  |  | ● |
| Perceived Stress |  |  | ● |
| Sadness |  |  | ● |
| Maternal Relationships |  |  | ● |
| Self-Efficacy |  |  | ● |
| General Life Satisfaction |  |  | ● |

<sup>1</sup> Administered to children ages 3-6 years old only;
<sup>2</sup> Condition 1 (Visual Scanning) and Condition 5 (Motor Speed) only are presented to children age 6-7 years old; all task conditions are presented to children $\geq 8$ years old (i.e., also including Condition 2: Number Sequencing; Condition 3: Letter Sequencing; Condition 4: Number-Letter Sequencing)
<sup>3</sup> Semantic categories only (not “First Letter”)
<sup>4</sup> Cognition Battery and Emotion Battery task versions selected based on subject age. The 4-Meter Gait Speed task is not age-validated for younger subjects but a raw score (time to complete task) is generated and used in our analyses.

If any information obtained during the above assessments raises concern that the child is at risk for harm due to suicidality, exposure to violence/abuse, or other reasons, the study team conducts a safety assessment and responds clinically as needed (e.g., referral for psychiatric evaluation), including referral to the Victim Support Unit at PCOE if necessary. Any care arising from the safety assessment and referral process is provided free of charge.

### Categorization of Cognitive Impairment

For our primary analysis of impairment we will use a Global Deficit Score approach ^23–26^. In a secondary analysis we will compare this approach to an approach defining cognitive impairment using Frascati criteria. ^27,3^ As an exploratory measure, we will determine which of these methods of classifying impairment has the highest correlation with “real-world” measures such as school performance and self-care as measured on the Brief Impairment Scale.

### Methods for assessing longitudinal cognitive trajectories

Utilizing the same population described above, the battery will be repeated annually for a total of two years of follow up for each subject. We will calculate a composite score (“NPZ8”) comprising performance across eight cognitive domains commonly affected in HIV: executive function/response inhibition, set shifting, attention, processing speed, immediate recall, verbal fluency, nonverbal reasoning, and motor speed. Test scores in each domain will be transformed for normality and converted into demographically-adjusted Z scores. The NPZ8 score is an age- and sex-adjusted mean of all of the z scores for the 8 domain scores. For the primary analysis, subjects will be assigned a Global Change Score based on change on the NPZ8 score over this period utilizing standardized regression-based (SRB) methods^30, 31^. SRB approaches involve the use of regression models to predict retest scores using formulae that account for baseline performance and demographic or other variables that may impact the rate of test–retest change. These normative SRB equations can then be used to determine whether the patient’s observed rate of change deviates significantly from expectation based on the change predicted from the model. In the SRB approach, subjects will be defined as having cognitive decline if the global change score is >2 standard deviations below the mean for HEU controls. We note that this approach will define relative decline compared to expected scores for age rather than absolute decline. In a secondary analysis, we will use group-based trajectory modeling (GBTM) to identify subjects with different developmental trajectories including cognitive decline ^32, 33^. GBTM is a statistical method for identifying developmental trajectories; the goal is to chart distinctive trajectories, understand what factors account for their distinctiveness and to test whether certain events modify a trajectory. The same methods will also be used to examine longitudinal trajectories of depression and anxiety scores.

### Statistical Methods

All statistical analyses will be performed using Stata (StataCorp 2018, *Stata Statistical Software: Release 14*, College Station, Tx, USA), with the exception of predictive modeling analyses detailed below for Aim 5 that will utilize relevant R packages. Rates of impairment will be compared between groups using logistic regression to control for confounders; and linear regression via generalized estimating equations will be used to summarize clustered domain and NPZ8 scores within individuals and to examine effects among groups, controlling for age, sex, socioeconomic status, and demographic and family characteristics. Risk factors, moderating factors, and mediating factors for impairment will be assessed utilizing latent variable methods such as Structural Equation Models (SEMs)^29^ and factor analysis. GBTM analyses will use the Stata “Traj” function that generates parameter estimates allowing the calculation of a) the probability of group membership; b) the predicted trajectory for each group; and, c) the posterior probabilities of group membership. A generalization of the GBTM model will be used to link baseline characteristics to the probability of group membership. Structural Equation Models (SEMs) using factor analysis for variable reduction will be built to assess the contribution of key factors to cognitive outcomes including 1) Biomarkers of immune activation (described further in Aim 3, below); 2) Clinical and demographic characteristics including age and sex; 3) Socioeconomic status; 4) Depression; and, 5) Family environment.

### Study Aim 3: Biomarkers of Cognitive Impairment and Cognitive Decline in HIV-Infected Youth in Zambia

For Study Aim 3, our goal is to identify inflammatory biomarkers and cell populations associated with cognitive impairment and with longitudinal cognitive trajectories as described in Aim 2 above. Approximately 8 ml of blood is taken from each subject at each annual study visit. To avoid confounding by effects of acute illness, subjects are screened for any recent illness or fever, and if present, phlebotomy is rescheduled for a time at least two weeks after resolution of their last illness.

### Cytokine Measurement

Samples for biomarker analysis are collected in 4 ml EDTA Vacutainer tubes by a trained study nurse or physician and brought to the lab on ice. Samples are then centrifuged using a horizontal rotor (swing-out head) for 15 minutes at 1200 g at room temperature, and plasma is collected with a transfer pipette and aliquoted into 200 μl aliquots in labeled cryovials, and stored in a −80° freezer until analysis. Each sample should thus have only a single freeze-thaw cycle. Samples are processed in batches using the Ella Multiplex Cytokine Analyzer according to the manufacturer’s instructions. The following biomarkers are measured in each plasma sample: TNF-α, sCD163, sCD14, CRP, IL-6 and IFN γ.

### Flow Cytometry Analysis

Protocols for our flow cytometry analysis have previously been described ^12, 13^. Briefly, samples will be drawn in ACD buffered vacutainers and taken for processing within 1 hour of blood draw. Analyses will be performed on a BD FacsVerse flow cytometer. Cell populations will be analyzed using Flowjo software. Samples will be stained with antibodies to CD14, CD16, CD61, and CD62P. For each sample, unstained cells are used to define the “Leukocyte” gate on a forward and side scatter chart (FSC/SSC). Cells in the leukocyte gate are used to formulate the next chart and subsequently, the monocytes are gated based on their FSC/SSC characteristics. These two gates are then applied to the cells stained with antibodies against CD14 and CD16 only. These cells are used to differentiate between two monocyte subtypes, CD14+ CD16− and CD14+ CD16+.The same cells are used to define the gate for detecting PMCs, i.e. monocytes expressing CD61, and are labeled as CD16− CD61+ and CD16+ CD61+. Each of these gates is then applied to the cells stained with all four antibodies to calculate percentages of CD16− and CD16+ PMCs in the total monocyte population. Unstained cells are used to define the gate comprised of platelets and leukocytes on a FSC/ SSC plot (labeled as “Cells” gate). Sizing beads (Mega Mix, Biocytex, Marseille, France) are also used to define the “platelets” gate (0.9–3 μ m) and to eliminate debris as well as microparticles. The cells stained with CD14 and CD16 antibodies alone are then used to define the CD61+ and CD62P+ gates, and these gates are then applied to the cells stained with anti-CD61 and anti-CD62P to calculate the percentage of CD62P+ platelets and CD62P expression as Median Fluorescence Intensity (MFI).

### Statistical Analysis

Plasma biomarkers and flow cytometry measurements will be log transformed or otherwise transformed for normality. For our primary analysis we will evaluate all biomarkers as continuous variables. If graphical evaluation indicates a clear threshold effect, biomarkers levels above the threshold will be evaluated as dichotomous variables. Correlations between biomarkers and flow cytometry measurements will be assessed using Spearman’s correlation coefficient. We will assess the association between each primary biomarker at baseline and Global Change Score using linear regression to control for confounders and explore the contribution of covariates including age, sex, socioeconomic status, and illness severity. Generalized linear mixed models will be used for longitudinal analyses relating cognition and biomarker levels across time. We will test for an interaction between HIV status, biomarker levels, and each cognitive outcome variable, and, if present, separate models will be fit for HIV+ subjects and the HEU control group. False discovery rate will be controlled for using an adjusted Bonferroni correction. Structural equation models with factor analysis for variable reduction will be utilized to investigate specific pathways (e.g. monocyte activation) and change in cognition over time.

### Study Aim 4: Neuroimaging Correlates of Cognitive Impairment in HIV-Infected Youth in Zambia

Brain MRI sequences are obtained using a 1.5T Siemens scanner located at UTH. No sedation or contrast agents are used. Sequences included: localizer scouts (3-plane); whole-brain sagittal three dimensional (SD) T1 weighted gradient echo sequence; T1, T2 and FLAIR sequences; DWI and ADC sequences; and Magnetic Resonance Angiography sequences. T1 sequences are optimized for volumetric assessments with a sagittal T1-weighted 3D turbo field echo sequence (echo time/repetition time = 5.12 ms/2200 ms; flip angle 15°; slice thickness 1 mm^2^; 256×256 acquisition matrix at 1 mm^2^) Each scan is anonymized and reviewed by a neuroradiologist with expertise in imaging in resource-limited settings, and who is blinded to subjects’ HIV status and clinical history. MRI scans are reviewed and annotated using the NeuroInterp database system.^22^ A clinical report is generated for each MRI, and verbal and written results are given to each subject by a neurologist. A second reader adjudicates any discrepancies between the Neuroradiologist MRI read and the neurologist interpretation of the scan.

### Image Processing

FreeSurfer (version 6.0.0, http://www.nmr.mgh.harvard.edu/freesurfer) is used to process structural T1-weighted MRI scans.^23-25^ The procedure includes skull-stripping, intensity normalization, Talairach transformation, segmentation of subcortical white matter and deep gray matter, and cortical gray/white matter boundary and pial surface reconstruction. Pial reconstruction was improved by the inclusion of T2 FLAIR sequences. FreeSurfer data processing was visually inspected for quality assurance and minor manual edits were made as necessary according to FreeSurfer documentation.

### Statistical Analysis

Before performing the primary analyses, descriptive statistics and graphical summaries will be obtained for each of the imaging measures to check for outliers and violations of model assumptions and to assess the need for transformations or non-parametric methods. We will also plot the primary measures as a function of age to examine the form of the developmental trajectories. In exploratory analyses we will examine the relationship between regional brain volumes and specific cognitive domains, e.g., correlation between executive function and frontal gray matter. We will also utilize the NeuroInterp output to construct heat maps according to global change score and brain volumes in order to identify features most associated with cognitive decline. For our primary analyses, we will test the association between baseline age-adjusted brain volumes (total cortical, gray matter, and white matter) and cognitive outcomes. Generalized Linear Mixed Models (GLMMs) will be used for longitudinal analyses relating imaging measures to NPZ8 change scores and biomarker levels across time. First, we will model developmental trajectories of brain measures, biomarkers, and cognition, including main effects of group (HIV+ vs. controls) and age, group by age interactions, and subject-level random effects. We will then add the neuroimaging measures as time-varying covariates. For all models we will examine the sensitivity of the results to medication type and other possible confounders or moderators (e.g., sex, depression, CD4 count, BMI). All statistical inference will be drawn from the marginal likelihood and is standard output from statistical software.

### Study Aim 5: Development of Predictive Model of Cognitive Decline in HIV-Infected Youth in Zambia

Study Aim 5 is a purely analytic aim. Using the clinical characteristics, biomarkers, and flow cytometry markers described above, we will construct predictive models for cognitive impairment and change in NPZ8 score. After developing preliminary models, we will evaluate the addition of imaging markers in the subset of subjects with MRI.

### Classification methods and feature selection

Individual features (e.g. clinical characteristics, biomarkers, imaging characteristics) will be used as input data to train several machine learning algorithms for binary classification and linear prediction. For each of the proposed methods, labels (to supervise the training of the classifiers) will be derived using both an SRB global change score approach and a GBTM approach, and models generated using each approach will be compared. We will study the performance of the following algorithms:

#### Logistic regression

We will initially perform feature selection via logistic regression, using principal component analysis for variable reduction for highly correlated variables. We will explore adopting a weighted L1-norm regularization, to penalize each feature (e.g. biomarkers, imaging) in the model according to its cost. Relevant R packages are glmnet.

#### Random forests and Penalized support vector machines (SVM)

Random forests is a powerful and flexible modeling technique typically used in classification. Random forests have the added value of automatic (data-driven) feature selection; only those variables used in defining the splits enter the model. In addition, random forests work well with medium-sized datasets. We will use the randomForest package in R^26^ for these models. SVM is a machine learning approach typically used in classification, and is based on the idea of finding a hyperplane that best divides a dataset into two classes. We will use the kernlab package in R for these models. SVM approaches are fairly robust to smaller datasets^71-73^, and using L1-norm regularization can easily incorporate the “cost” of each item into the model. Penalized SVM can utilize automatic feature selection, and is robust to correlated predictors ^49 51^. We thus hypothesize that a random forest or SVM approach will have superior performance characteristics compared to a logistic regression approach.

### Performance metrics and validation

To avoid overfitting and exploit our data to its fullest extent, we will perform 5-fold cross validation (CV) with ten repeats to compare the performance of the different classifiers and obtain reliable estimates of their sensitivity, specificity and accuracy. To assess predictive accuracy of the proposed models in the setting of unbalanced class distribution, our primary outcome measure will be the partial AUC-ROC for a specificity of 0.5-1. We will also consider additional metrics such as overall accuracy, F-measure, average recall, model complexity, and aggregate “cost” of the features deemed relevant to obtain an accurate prediction of decline.

### Validation

We will report the proportion of times that specific features were selected within subsamples of the data, and the accuracy of the classifier when limited to the particular subset of features selected in each CV fold. This way we will be able to carry out a fair assessment on the robustness and stability of the chosen features. In addition, we will report variable importance plots (VIPs) indicating the relative importance of each feature in the predicted response. For the L1 regularized methods such as logistic regression and SVMs, the magnitudes of the estimated (non-zero) regression coefficients are also indicative of variable importance. We will examine convergence between different ML methods and the extent to which features were independently selected as a further measure of internal validity. External validation of the models generated would be planned in a future study.

## DATA MANAGEMENT PLAN

All local data are entered into a password-protected Research Electronic Data Capture (REDCap)^27,28^ database installed on portable tablet computers and hosted at the University of Rochester. REDCap is a secure, web-based software platform designed to support data capture for research studies, providing 1) an intuitive interface for validated data capture; 2) audit trails for tracking data manipulation and export procedures; 3) automated export procedures for seamless data downloads to common statistical packages; and 4) procedures for data integration and interoperability with external sources. Data are collected offline in REDCap and then uploaded at least daily to a URMC-based server, permitting the PI to access and review data irrespective of location. The REDCap database is backed up daily and transferred to a secure file server accessible only to designated personnel; the server is also backed-up on a daily basis. All data and records generated during this study will be kept confidential in accordance with Institutional policies. De-identified data will be shared with selected collaborators (this is disclosed in the informed consent process/forms) and stored for use as a control dataset for future studies. After publication of primary results, de-identified data will also be posted in secure servers (e.g. BioRxiv) along with analytic code in order to increase data transparency.

## SAMPLE SIZE ESTIMATION

Full details of the statistical approach for the respective study aims will be provided in forthcoming publications. A sample size of 200 subjects per group was initially planned based on simulation studies for each aim, with the goal of ensuring model stability and avoiding overfitting for regression models containing up to 10 independent variables, and for structural equation models containing up to ten latent variables. The least power would be available to evaluate dichotomous outcomes (e.g. cognitive impairment) within subgroups. Assuming that rates of cognitive impairment were at least 20% in the HIV-infected population, this sample size would allow model stability for evaluation of up to 4 independent variables as risk factors for cognitive impairment, with >80% power to detect odds ratios of 1.5 or greater. Study Aim 4 (Neuroimaging sub-study) is powered to detect a difference of at least 5% in total and regional brain volume between HIV-infected and HEU groups. The expansion of the sample size to 300 subjects per group was determined in order to ensure sufficient subjects in each age strata at the lower end of the age spectrum in order to investigate an interaction between age and cognition.

## CHALLENGES, SOLUTIONS

### English-Language Fluency and Literacy, and Numeracy in School-Age Children in Zambia

#### Challenge

As noted earlier, English is the official language of Zambia. We expected all subjects initially enrolled in the study (ages 8 years and older) to have had at least several years of English language exposure in school. All children attending school receive instruction in English; from Grade 1-4, familiar/local languages are used as a medium of instruction in content areas while English is used as a medium of instruction from Grade 5 to the Tertiary level.^29^ We discovered soon after beginning enrollment that some children and adolescents with regular school attendance still struggled to understand cognitive test instructions in English.

Poor literacy and numeracy^30,31^ was also evident on the DKEFS Trail Making Test, for which two sub-tasks require rapid, accurate sequencing of numbers (from 1-16) and the alphabet (letters A-P) in order, respectively; a third sub-task (Number-Letter Switching) requires rapid alternation in the sequencing of numbers *and* letters (e.g., A-1, B-2, etc…). This third component of DKEFS TMT was pre-selected as one of our measures of executive function (i.e., set-shifting), but in some subjects, we have found that poor performance on this task (i.e., completed with errors, or too slowly) may not be due to a specific impairment in set-shifting but rather due to lack of familiarity with core alphabetic or numeric principles.

#### Solutions

We have instituted several solutions. First, test instructions for all NIHTB-CB and neuropsychological assessments have been translated/back-translated by local staff into Chinyanja, the vernacular language throughout the Lusaka community. Test stimuli were not translated, as this would have necessitated re-validation of the measures and establishment of vocabulary (in Chinyanja) of similar frequency and difficulty level as in English. When subjects present for study visits, local study staff ascertain their ability to converse and understand task instructions in English vs. Chinyanja, and proceed accordingly with the cognitive assessments (NIHTB-CB and neuropsychological battery). We added a field to our data collection forms to indicate whether task instructions were presented in English or in Chinyanja.

Second, to address concerns related to limited literacy and numeracy skills, we added a brief screening at the beginning of the neuropsychological assessment battery, to guide interpretation of the DKEFS TMT results. In particular, we now ask all subjects to read and write (to dictation) letters and numbers in English, and recite the English alphabet and numbers in order. The tasks are presented in the same way for each subject and responses are recorded on a standard data collection sheet. We recognize that psychometric properties (reliability, validity) of the task have not yet been established for this population. Finally, we removed a planned task, the “First Letter” task of the NEPSY Verbal Fluency test; this test involves rapid naming of words beginning with a specific first letter of the English alphabet and depends upon mastery of basic phonemic awareness (understanding of letter-sound correspondences).

### Age of Onset of Neurocognitive Impairments in HIV-Infected Children

#### Challenge

Preliminary data (*unpublished)* from the first year of the study suggested that by age 8 years old, there were already substantial differences in cognitive test performance between the subjects with HIV and the HEU group, and that these differences increased over the enrolled age range (HEU group performance outstripped HIV-positive group). Consequently, we lacked information on possible earlier characteristics or predictors of neurocognitive deficits in this group of perinatally infected children. It was also apparent after the first year of successful recruitment that we had established the largest existing dataset of development and cognition in children and adolescents with and without HIV infection in this age range in Zambia. However, we were aware of other studies taking place in Zambia of younger (than our sample) children with HIV infection, and realized there was a potential missed opportunity to optimize comparability of datasets.

#### Solution

Beginning in Year 2 of the study, we have expanded recruitment to children from 3-7 years of age to permit evaluation of potential earlier onset of cognitive impairments, and to characterize the trajectory associated predictors of cognitive impairment in pre-school age children with HIV infection compared to the HEU control group. The addition of the younger age group also provides an opportunity to collaborate with another study of children with HIV-infection in Zambia, the Cohort of HIV-Associated Seizures and Epilepsy in Zambia study (“CHASE”; PI: Birbeck), to address hypotheses of common interest. In the CHASE study, children with new onset seizures are prospectively enrolled, an etiological assessment is performed, and seizure and developmental outcomes are assessed. The neurocognitive assessment battery selected for this younger age group in HANDZ includes overlap with the CHASE protocol, particularly the use of the MDAT^15^ in children age 3-6, and the UNIT-2^14^ in children age 6-7 years old. Use of the UNIT-2 in children age 6-7 years old also allows continuity of measurement in the HANDZ study, where this assessment tool is also used in all other subjects, age ≥ 8 years old. Selected data from the HANDZ sample of children age 3-7 will contribute to all of our original study aims with the exception of development and validation of the ZDAP.

### Cultural Boundedness of Test Stimuli

#### Challenge

Several tasks were selected for apparent minimal cultural influences, but were subsequently found to be not culturally relevant for children in Zambia, due to inclusion of unfamiliar vocabulary (e.g., certain foods or animals not known to Zambian youth). In particular, we had selected the Peabody Picture Vocabulary Test-Fourth edition (PPVT-4)^32^ as a simple test of receptive vocabulary within the neuropsychological assessment battery (Aim 2) and to aid in validation of an analog measure from the NIHTB-CB (Picture Vocabulary Test; Aim 1). There were more items than expected that subjects reported lack of familiarity with, even when translated terms were provided informally Chinyanja by study staff, to assess whether difficulty was due to a language barrier or unfamiliarity with the underlying concepts. A related, unexpected challenge was that children also had difficulty understanding some instructions (and verbal stimuli) presented on the NIHTB-CB because of the American-accented recorded voice used with the test.

#### Solution

We removed the PPVT-4 and NIHTB-CB Picture Vocabulary Test from their respective test batteries soon after the study was initiated. A Zambia-adapted version of the PPVT was considered but deemed not suitable for our main sample of children ≥ 8 years old, as it was developed with item content only suitable for younger children ≤ 6 years old.^33^ This would have led to ceiling effects with our sample which enrolls adolescents as old as 19 years of age. Where feasible for NIHTB-CB tasks (when it would not interfere with task performance), instructions and task stimuli were re-stated by the examiner using Zambian-accented English; instructions could also be presented in Chinyanja if necessary, but we did not translate task stimuli themselves due to similar concerns as with the PPVT-4.

### Summary

The HANDZ study has 5 key aims, including 1) Developing and validating instruments for the assessment of neuropsychological function in Zambia 2) Identifying the incidence, prevalence, and risk factors for neuropsychological disorders in Zambian youth with HIV 3) Identifying biomarkers for neuropsychological disorders in HIV-infected youth Zambia 4) Identifying neuroimaging correlates of neuropsychological disorders in HIV-infected youth and 5) Developing predictive models for longitudinal cognitive and psychiatric outcomes. The HANDZ study will attempt to fill several key knowledge gaps related to neuropsychological disorders in children and adolescents with HIV, including lack of longitudinal outcome data in older children and adolescents with HIV in resource-limited settings. The primary innovation of this study includes use of multiple modalities to assess cognitive function and use of modeling techniques for prediction of longitudinal outcomes. In addition, this study will allow determination of the relative importance of multiple factors potentially contributing to neuropsychiatric disorders in youth with HIV, setting the stage for future clinical trials to prevent or treat these disorders.

## Data Availability

This is a methods paper and as such there are no data/results reported in this paper.

## Acknowledgments

We gratefully acknowledge the efforts of the local staff in Lusaka, Zambia: Beauty Matoka, Ivy Makulu, Margaret Mbulo, Milimo Mweemba, Priscilla Mweemba.

